# When is SARS-CoV-2 in your shopping list?

**DOI:** 10.1101/2020.06.11.20128850

**Authors:** Gustavo Hernandez-Mejia, Esteban A. Hernandez-Vargas

## Abstract

The pandemic of coronavirus disease 2019 (COVID-19) has caused, by May 24th 2020, more than 5.3 million confirmed cases worldwide. The necessity of keeping open and accessible public commercial establishments such as supermarkets or pharmacies increases during the pandemic provided that distancing rules and crowd control are satisfied.

Herein, using agent-based models, we explore the potential spread of the novel SARS-CoV-2 considering the case of a small size supermarket. For diverse distancing rules and number of simultaneous users (customers), we question flexible and limited movement policies, guiding the flow and interactions of users in place. Results indicate that a guided, limited in movement and well-organized policy combined with a distance rule of at least 1 m between users and a small number of them (15) may aid in the mitigation of potential new contagions in more than 90% compared to the usual policy of flexible movement with more users (30) which may reach up to 64% of mitigation of potential new infections under the same distancing conditions. This study may guide novel strategies for the mitigation of the current COVID-19 pandemic, at any stage, and prevention of future outbreaks of SARS-CoV-2 or related viruses.

## Introduction

The world is currently impacted by the coronavirus disease 2019 (COVID-19) pandemic which is caused by the severe acute respiratory syndrome coronavirus 2 (SARS-CoV-2) that, by May 24th 2020, has caused more than 5.3 million confirmed cases. Many authorities at the global level have adopted measures known as “physical distancing” or “social distancing” (SD) which go from schools and workplaces closure to limiting gatherings, quarantining infected individuals and their family members and self-quarantining to avoid getting infected^1,2^. Besides, hygiene measures have been large recommended, these include washing hands, covering up when coughing or sneezing, and avoid touching the face with unwashed hands since these may be frequently touched by other potentially infected people. One of the first reported studies of the potential effect of SD strategies on COVID-19 burden was developed in Singapore^3^ and the interventions continued in other several European countries^4^. The objective of the SD interventions is to decelerate the spread of infection and reduce the intensity of the epidemic to avoid potential overwhelming health systems and, at the same time, gain a time-line to develop treatments and vaccines. However, a matter of consideration is that these interventions may hold for long periods with certain relaxing phases as population immunity gradually increases allowing the measures to loosen^1,5^. There is still limited evidence to support SD measures as schemes of reducing transmission and slowing down the spread, however, accessible evidence may suggest that staggered and cumulative implementation of these interventions, in conjunction with testing and contact tracing of all suspected cases^6^ following close scientific and ethical basis, show the most effective outcome against COVID-19^7–9^.

One of the most important players of goods supply chains for the population, as well as key social encounter places, are public commercial establishments. Supermarkets, pharmacies, grocery stores, discount stores, and other commercial chains can act as a route of spread for both, clients (users) and place workers^10^. Public commercial establishments are generally enclosed places usually with one entrance and one exit gates that under normal situations, SD measures and the number of users has no relevance. Inside the establishment, users freely interact with each other following pretty well-defined trajectories as those of supermarket corridors. In this manner, users and working personnel in place can be characterized in terms of their movements when following the corridors as well as their physical interactions and the physical distance between and among them, under a physical metric. In this direction, agent-based (AB) modeling schemes can map all desired characteristics of users and workers, the agents, and their interactions in the establishment’s corridors, identified as agents’ trajectories or paths. AB schemes have previously been used to devise strategies on health behaviors, social epidemiology^11^, and to advise on the mitigation of previous influenza outbreaks^12^ as well as to explore immune responses to this virus^13^. Under this approach, AB schemes are also to be employed for testing population strategies for enclosed environments for the coronavirus pandemic^14^ that may add to the forces of non-pharmaceutical interventions^15^ especially in high-density population places, like urban scenarios^16^. Moreover, since specific conditions and characterizations can be implemented for agents and trajectories, diverse behavioral aspects can be designed and tested, for instance, the checkout area design of a supermarket to model the users flow and guidance strategies^17^.

Herein, we explore the potential spread of the novel SARS-CoV-2 in public commercial establishments using an AB approach. We analyze diverse distancing rules between users and different population sizes in a layout that represents a small-to-medium size supermarket. Also, we test flexible and limited movement policies that guide the flow and interactions of users inside the supermarket. Results indicate that a guided, limited in movement and well-organized policy may significantly collaborate into the mitigation of potential new contagions of SARS-CoV-2 in public commercial establishments.

## Methods

### Commercial establishment layout

We consider a general layout of a public commercial establishment, in this case, a small-to-medium size supermarket structure. The considered dimensions of the establishment may also be suitable for pharmacies, discount markets, and convenience stores.

Figure 1 depicts the considered structure, accounting for one entrance access and one exit. Corridors and shelves are distributed within the supermarket in such a manner that the users can freely move through them, avoiding bottlenecks. Figure 1-A shows the corridors marked with dot-colored lines and an identifier number. The marks stand for the distancing rule of an agent when moving, although it is also allowed that more than one agent can stand in the same mark as they freely move. All paths are designed following this metric to map all the supermarket corridors. Figure 1-B shows the directions that agents can follow on each corridor highlighting the routes available when moving on the layout. Whenever an agent reaches the end of a corridor and can take more than one corridor to continue, it selects the next one based on a probability of selection (50%) with a binomial distribution. We consider three checkout stations operating with one supermarket worker (cashier) per station. As shown in Figure 1, the labels on the shelves indicate a general conception of the supermarket areas, these can naturally be named differently. The model is rendered in a Cartesian layout where each agent is represented through its *x*- and *y*-coordinates that change as the agent moves through the supermarket paths.

**Figure 1.**
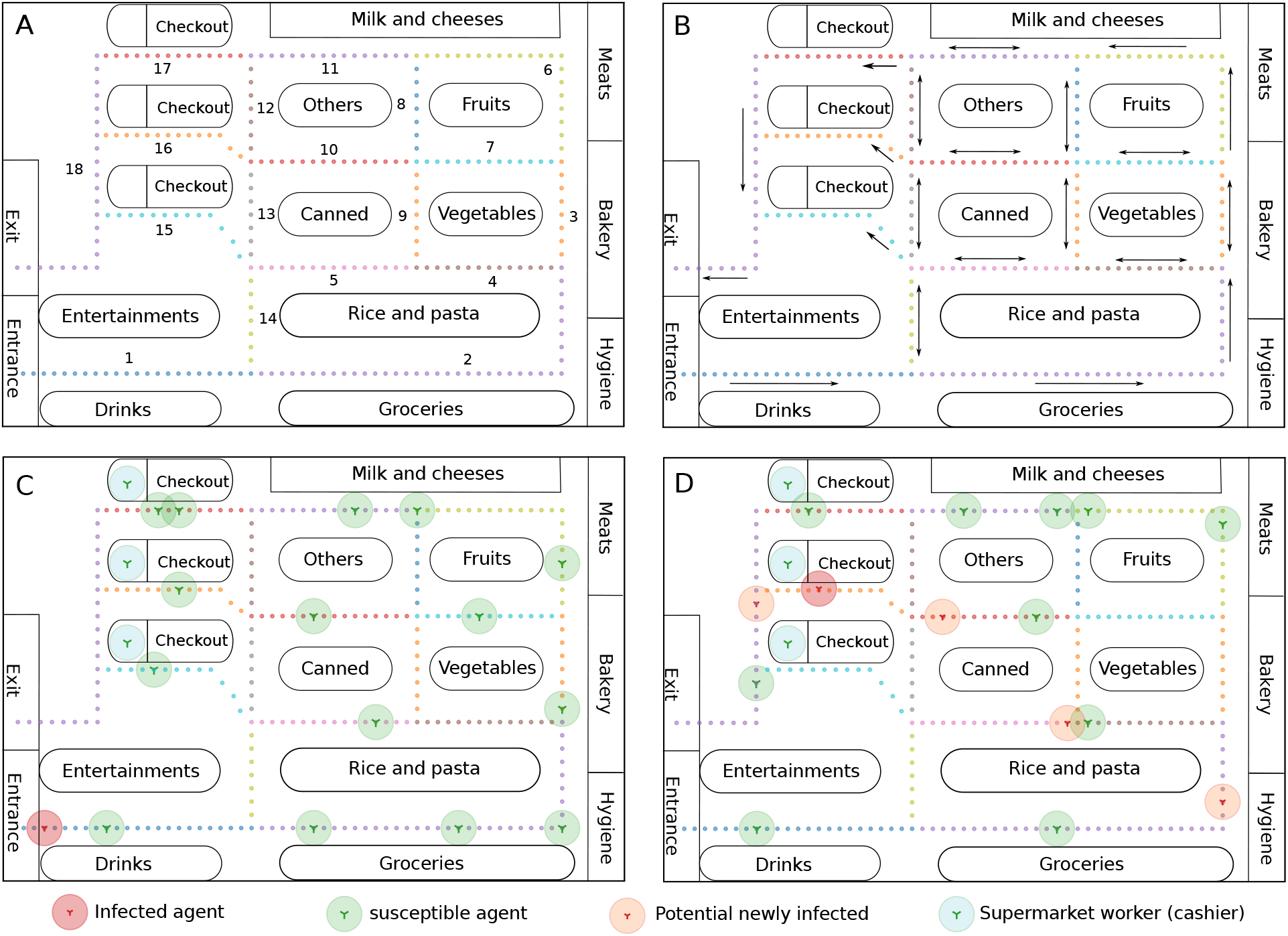
Supermarket layout and spread model. A) Corridors in the layout (paths) are highlighted with dot-colored (marks) lines, showing the routes that agents can follow, also, the identifier number of each corridor is shown. The layout represents a small to medium size supermarket of 30 × 16 m. In this case example, the distance between marks is 50 cm (distancing rule). Since this is a general layout, the labels of the areas of the supermarket can be modified for a more suitable representation. B) Supermarket layout with arrows indicating the directions that each path can handle, some of them are bidirectional, some others unidirectional. C) Agents are divided into four subsets, susceptible (green), potential newly infected agents (orange), workers of the supermarket (cashiers, blue), and the infected agent (red). All agents, except the cashiers, can freely move all around the supermarket using and standing over the marks of the paths, therefore, the distance between agents is given by the separation of the marks of the paths. The layout shows the initial allocation of all agents considering that an infected agent enters the supermarket and moves around it following the paths. D) The infected agent is now in the checkout area after some movements in the layout, four potential newly infected agents have been produced.

### Spread model

The model considers the uninfected (*U*) and infected (*I*) populations, represented by agents *U*_*j*_ and *I*_*i*_, respectively. We consider a potential contagion from an infectious agent to have a probability of spread (*P*_*Spread*_) of 50% with a binomial distribution. The spread is also governed by the “physical” distance (Euclidean distance, *EUC*(*I*_*i*_,*U*_*j*_)) between the infectious agent *I*_*i*_ and the uninfected one *U*_*j*_. A potential contagion must therefore first satisfy a minimum distance threshold (*M*_*D*_) between agents and a positive outcome of the binomial probability. The simulation initiates with a fixed population size *N* = *U* + *I* and, each potential newly infected agent is taken from the *U*-population and appended to the *I*-population.

A case example with an initial allocation of 15 uninfected users, three cashiers and one infected user is shown in Figure 1-C. This presents a case in which the infected agent (user) enters the supermarket and starts moving using the paths following the directions of Figure 1-B, all other agents also move and can leave the supermarket. Those agents that leave the supermarket, infected or uninfected, are replaced by uninfected agents. After some movements inside the supermarket, the infected agent is on the checkout area and has had potential contagion with at least four other uninfected agents, as shown in Figure 1-D. A case example with an initial allocation of 15 uninfected users, three cashiers and one infected user is shown in Figure 1-C. This presents a case in which the infected agent (user) enters the supermarket and starts moving using the paths following the directions of Figure 1-B, all other agents also move and can leave the supermarket. Those agents that leave the supermarket, infected or uninfected, are replaced by uninfected agents. After some movements inside the supermarket, the infected agent is on the checkout area and has had potential contagion with at least four other uninfected agents, as shown in Figure 1-D. Finally, the simulation steps are as follow: all agents move from the initial allocation to the immediate next one (agent steps) according to the distancing rule and the corridor direction, the minimum distance threshold and contagion probability are checked (all uninfected agents respect to the infectious), potential newly infected are generated if applicable, and all agents move again. The simulation stops when the infectious agent reaches a maximum number of agent steps or leaves the supermarket. The maximum number of steps also allows representing the spent time in the supermarket this can go from 15 to 40 minutes.

## Results

### Flexible movement policy

We aim to test the impact of different distancing rules and population sizes on the generation of potential newly infected. Therefore, we test 5 distancing rules, 30 cm, 50 cm, 1 m, 1.5 m, and 2 m. Besides, we test populations of 15, 20, 30, and 50 uninfected agents. In all cases, we account for 3 cashiers and one infected agent. The minimum distance threshold is 1.5 m. We perform 1000 simulations per case registering the number of potential newly infected cases as well as the percentage of repetition (frequency) of each case for all the distancing rules and population sizes. We identify this test as the flexible movement policy, as all agents can move using the directions of Figure 1-B with almost no movement limitations. The results with the number of potential newly infected from all simulation cases and the total of potential newly infected are depicted in Figure 2.

**Figure 2.**
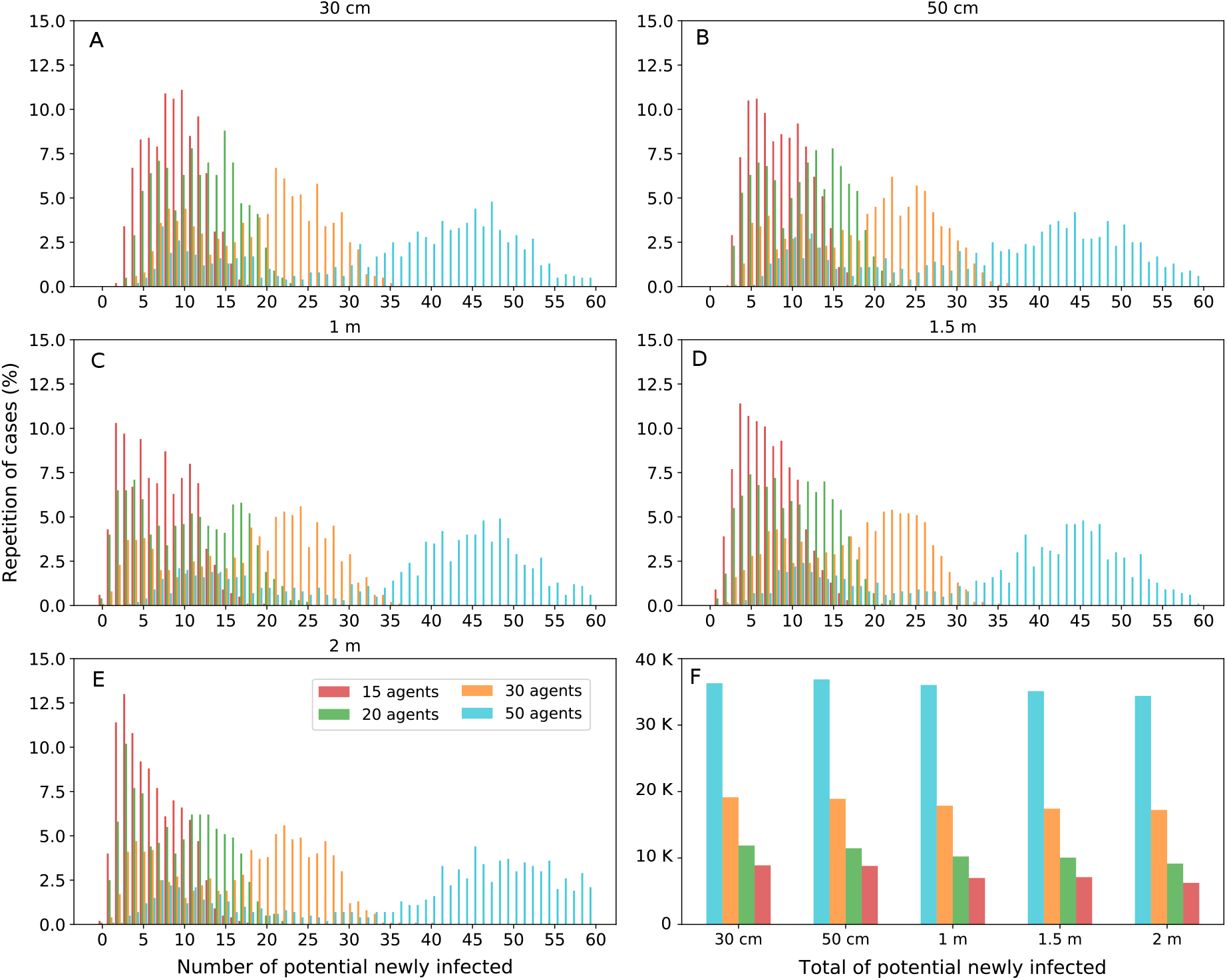
Flexible movement policy with potential newly infected agents considering different distancing rules and population sizes. Each panel presents the distribution of the number of potential newly infected agents and the percentage of case repetition for populations of 15 (red) 20 (green) 30 (yellow) and 50 (blue) uninfected agents and three cashiers. A-E) The distancing rules are indicated on top of panels. Each distribution case reports the results of 1000 simulation. F) The total of the potential newly infected agents for each case of population size and distancing rules.

The distribution of the percentage of repetition of cases greatly varies with respect to the number of potential newly infected agents as the population size increases. The distribution of cases considering 15 and 20 uninfected agents, the red and green histograms, show similar behavior in all distancing rules. Using the rules of 30 cm to 1 m (Figure 2-A-C), for instance, the shape of the histograms show a uniform tendency while for rules of 1.5 and 2 m both histograms present a higher percentage of repeated cases for up to 5 newly infected. However, this is not the case when comparing with the 30 agents histogram (yellow), in which all cases present a bimodal distribution with peaks around 5 to 10 and 20 to 25 newly infected agents. This behavior may be reflecting the separation of cases in which the infectious agent spends less time in the supermarket compared to those cases spending more time since the interaction with other agents is also being reduced. Similarly, the distribution of the 50 agent histograms presents two peaks in all distancing rules, the first shape peaking from 5 to 15 newly infected and the second shape peaking from 45 to 50 newly infected agents.

The distancing rule may weakly affect the total of potential newly infected, however, the smaller uninfected populations may add to the mitigation of potential contagions. In Figure 2-F, the total number of potential contagions present slight differences between distancing rules for populations of up to 20 agents, both cases with around 10 thousand (10 K) total cases. In the case of 30 agents, the total of potential contagions remains around 20 K in all distancing rules, and, for 50 agents, the total reaches almost 40 K of potential newly infected. The greater difference in the number of potential newly infected agents relies on keeping the population inside the supermarket to be less than 30 users, preferably between 15 and 20 users.

### Limited movement policy

We further test a limited movement policy aiming to explore the impact on the potential contagions mitigation. The distancing rules remain as in the flexible movement policy, however, the corridors’ usage directions and some access to corridors are limited. Figure 3-A shows the framework of the limited movement policy in which the identifier number of each corridor remains as those in Figure 1-A, however, the access to corridor 14, for instance, is fully restricted. There are some corridors in which a user can enter but not exit to the same corridor, for instance, corridor 7 can be accessed from corridor 3 but can not return to corridor 3 or 6. If corridor 7 is accessed from corridors 8 or 9, the user must return to use one of these corridors. These rules are indicated by the oval arrows and elbow arrows in Figure 3-A and pretend to guide users’ flow.

**Figure 3.**
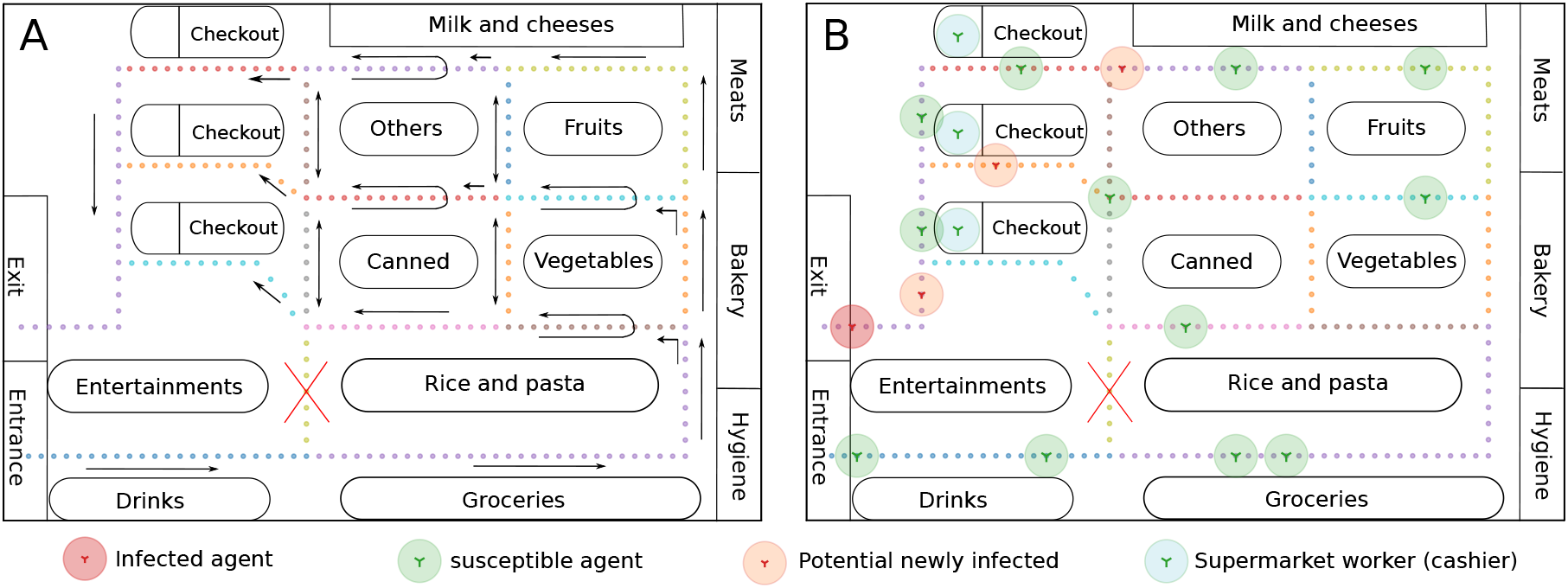
Limited movement policy layout. A) The corridors conserve the same identification number as in Figure 1-A as well as the dotted lines indicating the distancing rule of the paths. The corridors also show the new movement policy highlighted by oval, elbow, unidirectional and bidirectional arrows all over the supermarket layout. Some corridors like the number 8 and 9, for instance, remain in a bidirectional policy while others like 4 and 7 now present an oval arrow policy. The last indicates that these corridors can be, for example, accessed from corridor 3 but no return can be made to this corridor. Also, corridors 4 and 7 can be accessed from corridors 8 and 9, and users must return to use one of these corridors. Lastly, corridor 14 is no longer available. B) The last steps of a case example simulation accounting for 15 susceptible agents and the limited movement policy, the initial allocation of agents is similar to the one in Figure 1-C. In this case, the newly infected agents remain closer to the infectious agent, in the checkout area, since agents follow the guidance rules.

The general behavior depicts that accounting for the same initial users’ allocation shown in Figure 1-C, users interact with a limited number of other users, mostly with those who entered the supermarket. A case example is depicted in Figure 3-B where the infectious agent leaves the supermarket and produced three potential newly infected agents that also are about to leave the supermarket. No other newly infected agent is found out of the checkout area and those agents in the remaining areas are all susceptible.

Results of the limited movement policy are shown in Figure 4 accounting for the same distancing rules and population sizes as in the flexible movement framework. Using the limited movement policy, the distribution shape of potential newly infected agents shows to follow a gamma distribution shape when exploring up to 30 susceptible agents for all of the distancing rules, generally peaking from 0 to 5 potential new contagions when using up to 20 susceptible agents. The distribution of cases using 30 agents peaks from 0 to 10 potential new contagions. The frequency of cases also reached 10% more repetition of cases for the tests with populations of 15 and 20 agents, compared to the flexible movement test. For the case of 50 susceptible agents, the distribution shape is mostly uniform with less than 10% of repetition cases reported. Importantly, the total of potential newly infected remains in less than 10 K in all distancing rules for the tests with 15 and 20 agents. For the case of 30 agents, the total potential contagions are above 10 K for the distancing rules of 30 and 50 cm, however, potential contagions are below 10 K when the distancing rule is of at least 1 m. Finally, the case with 50 agents shows to produce more than 20 K potential new contagions in all distancing rules.

**Figure 4.**
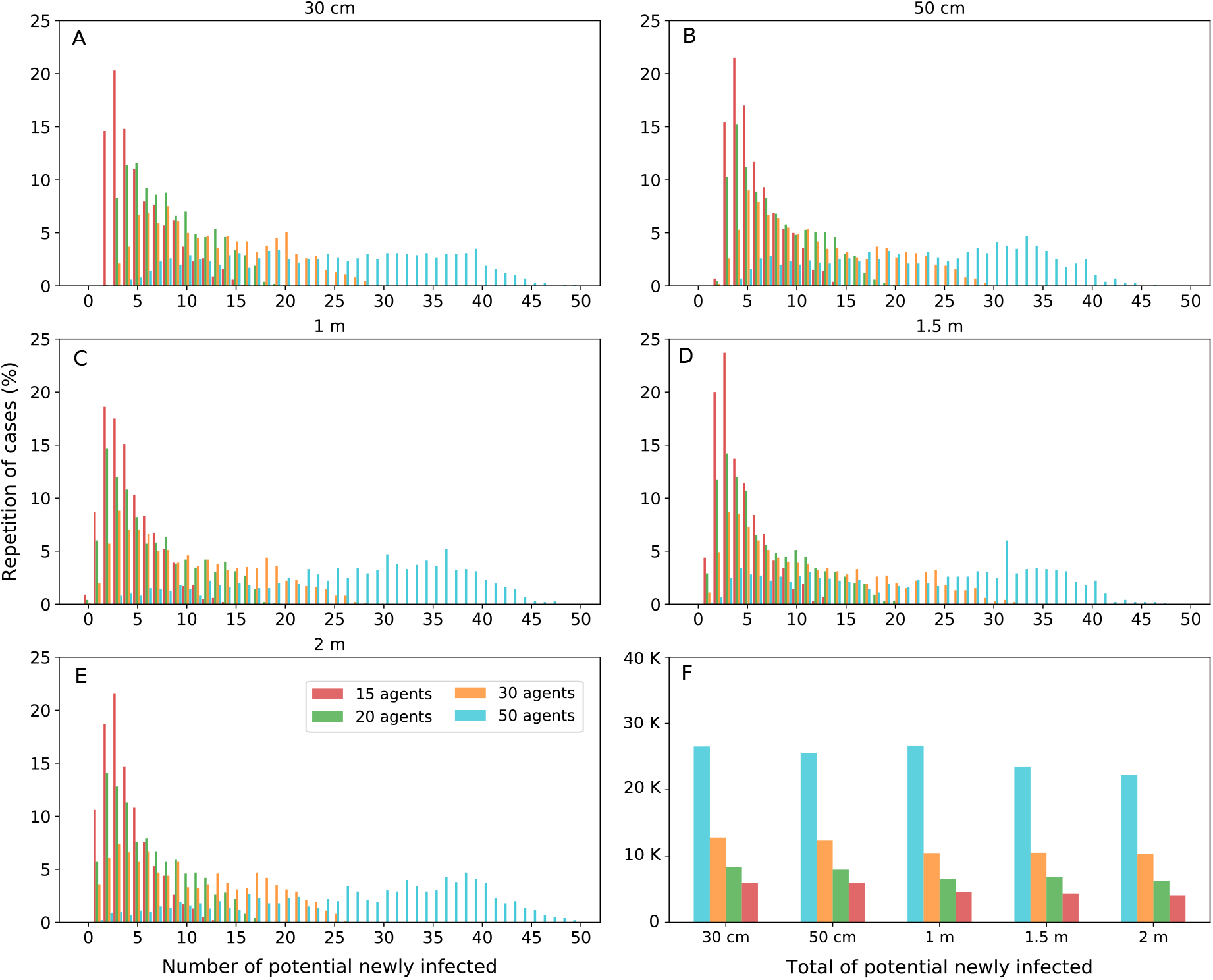
Limited movement policy results. The number of potential newly infected agents and case repetition percentages for different population sizes considering distancing rules of A) 30 cm, B) 50 cm, C) 1 m, D) 1.5 m, and E) 2 m. Each distribution case reports the results of 1000 simulations with populations of 15 (red), 20 (green), 30 (yellow), and 50 (blue) uninfected agents, and three cashiers. F) The total of the potential newly infected agents for each case of population size and distancing rules with the limited movement approach.

For a clearer comparison and appreciation of the tested policies we show in Figure 5 the histograms of both, flexible and limited movement policies. The histograms report the total of potential contagions for all population sizes and distancing rules. Considering the flexible policy, the ideal distancing rule may be at least 1.5 m between users and up to 15 users at a time. However, when using the limited movement framework, the number of users can be increased to 20. A similar panorama can be observed for 30 users since the distancing rules do not present great variation among them in the total of potential contagions, however, contagion mitigation can benefit from the limited movement approach since the total number of potential contagions can be reduced for up to 10 K compared to the flexible movement policy, combined with a distancing rule of at least 1.5 m. In a similar direction, the maximum number of possible contagions is observed when 50 users simultaneously interact in the establishment, with a similar impact of the limited movement policies on the potential new contagions, respect to the 30 users case. On average, up to 10 K fewer cases can be reached with the limited movement policy, compared to the flexible movement approach. However, the size of the population may prevent the benefit from further distancing rules and movement policies.

**Figure 5.**
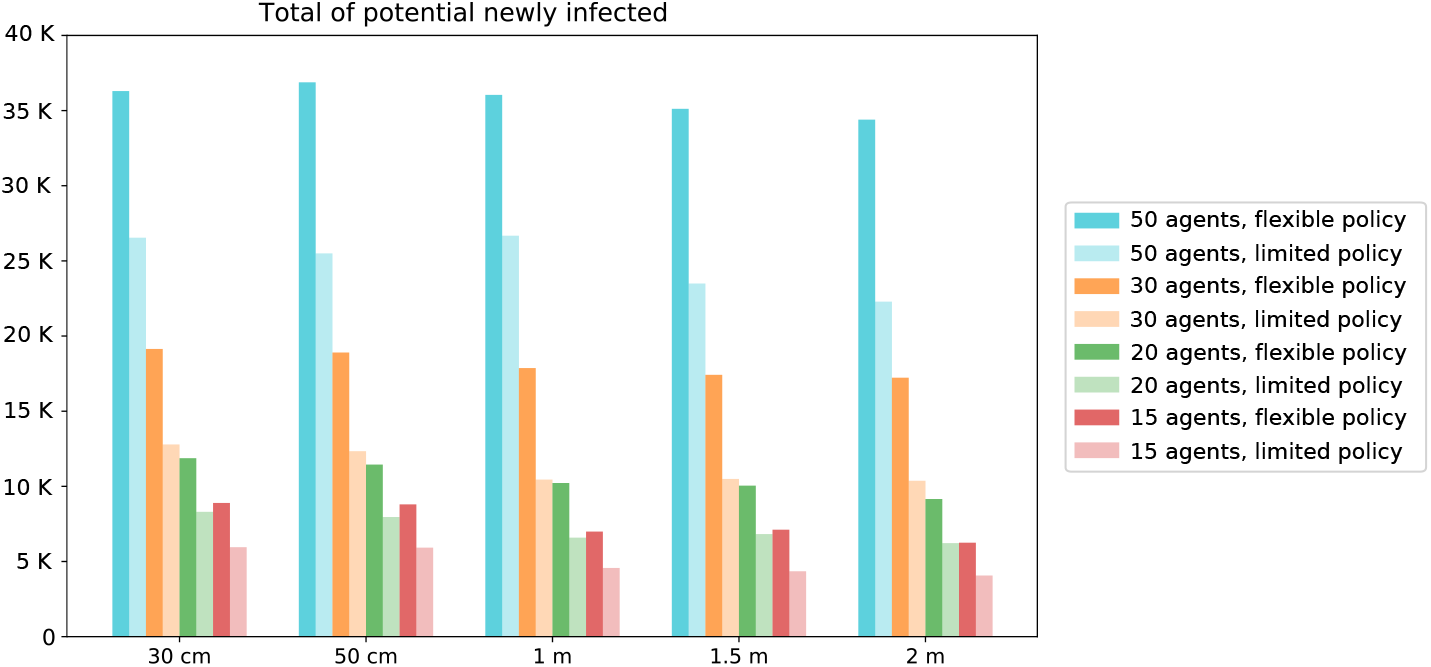
Comparison of flexible and limited movement policies. The histograms show the total number of potential newly infected for all population sizes and distancing rules in the test. The flexible movement policy histograms are depicted with darker colors, reporting the data of Figure 2-F. The limited movement framework is depicted with lighter color histograms and report data of Figure 4-F.

We further analyze the impact of the tested schemes on reducing the viral spread through the percentage of mitigation of potential contagions, as reported in Table 1. In this approach, we consider that a population of 50 users is to enter the supermarket, however, we explore different cases using the complete population or subsets of it to enter the supermarket at a time. Accounting for the simulations results in Figure 5, we compute the mitigation benefit as the percentage of users that remain susceptible after each of the performed simulations for both policies, flexible and limited movement. The distancing rules and the fraction of users (number of users) remain as in Figure 5. The number of users remains constant as those that leave the supermarket, potentially infected or not, are replaced by susceptible users.

**Table 1.**
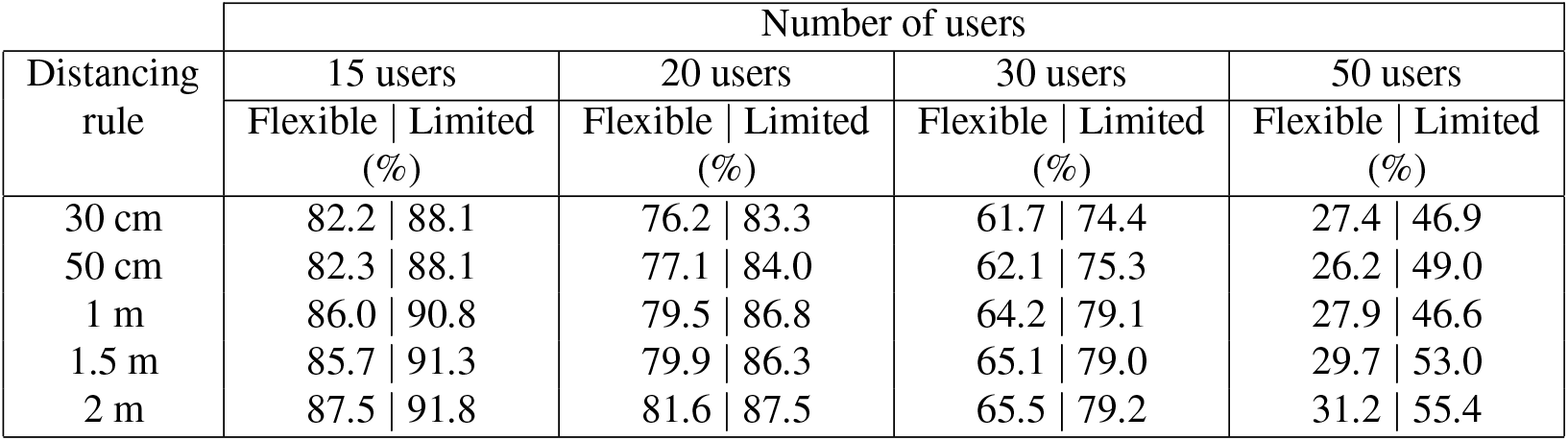
Percentage of mitigation of potential contagions. The percentages are presented according to the distancing rules and number of simultaneous users of the commercial establishment. A population size of 50 potential users is considered to evaluate how the number of simultaneous users, distancing rules, and movement policies impact the percentage of mitigation, which is the portion of users that remain susceptible after a simulation case. The percentages of mitigation are presented for the policies of flexible movement and limited movement.

The lowest percentage of mitigation of potential contagions is observed, as expected, whenever the complete population (50 users) accumulates inside the establishment for the distancing rule of 30 cm with the flexible movement policy, with only 27.4% of potential spread mitigation. Since the establishment is considered to be crowded, the distancing rules may have a negligible impact on the spread mitigation, reaching a mild 31.2% of potential mitigation with 2 m distancing. On the other hand, if the limited movement policy is considered, the mitigation of potential spreads reaches more than 50% for distancing rules of at least 1.5 m. A resemblance of the total population case is observed if using 30 users at a time, for the flexible movement approach, for instance, the percentage of mitigation of contagions reaches 65.5% for a rule of 2 m distancing, while the limited movement policy presents 79.2 % of potential mitigation for the same distancing rule. Of note, using the limited movement policy with rules of at least 1 m distance, the potential mitigation can reach almost 80%, a promising scheme compared to the limitations of the flexible approach that presents up to 65% of potential mitigation.

For the case of 20 simultaneous users, while the flexible movement policy reaches maximum potential mitigation of 81.6%, naturally with the 2 m rule, the mitigation using the limited movement approach reaches up to 87.5% for the same distancing rule. In this direction, whenever up to 15 users are allowed to share the establishment, the mitigation of potential new contagions benefits from both movement approaches and distancing rules, as may be expected. However, clearer differences can be found when comparing movement policies. In the common approach which is represented by the flexible policy, the percentage of mitigation reports at least 82% and reaches 87.5% for 2 m distancing, while the limited movement policy already reaches 88% of potential mitigation for the 30 cm policy. The reason for this outcome is that the infectious agent may remain in closer contact with a limited number of users when the movements are restricted, regardless of the distancing rule. Moreover, if a limited movement approach is performed in combination with at least 1 m of distancing rule, the mitigation of potential contagions reaches more than 90%, guiding potential schemes of viral spread mitigation.

## Discussion

Considering the tested policies, it is clear that to mitigate the spread of the virus and therefore limiting potential new contagions, commercial establishments can set distancing rules between users but also limit the number of users inside the establishment at a time. We depict the case example of a supermarket testing different levels of distancing rules and the number of users, besides, we explore two motion policies, the flexible and limited movement policies, whose results comparison are shown in Figure 5. Taking these results, we estimate the percentage of mitigation of potential new contagions for the flexible and limited movement schemes, accounting for the number of users and distancing rules. Results are reported in Table 1. Under this approach, if a percentage of mitigation of at least 85% is desired to reach, the flexible movement policy may be successful allowing the minimum number of users, 15 users, and a distancing rule of more than 1.5 m between users (86%). On the other hand, whenever the limited movement policy is used the number of users should be up to 20 and keep a distancing rule of at least 1 m (86.8%). Moreover, if more than 90% mitigation of potential viral spread is the target, then the limited movement policy with at least 1 m distance rule should be used. A target of more than 90% mitigation would not be overcome using the flexible movement approach. Of note, the percentage of mitigation scheme support not only the distancing rules and number of users schemes but also guided frameworks that allow users to access all areas of commercial establishments, in a well-organized manner, while significantly reduce the potential of SARS-CoV-2 contagion at any stage of the pandemic. The guided-limited movement policies may be also implemented in outbreak events of similar viruses, such as influenza A virus.

Hygiene measures, social distancing, gathering prevention and self-isolation are some of the most common recommendations that governments and authorities have indicated to combat the current pandemic, however, policies that add to viral spread mitigation in commercial establishments are still to be better implemented. A limitation of these kinds of policies may be the implementation itself, since they can represent extra work and pressure to the personnel establishments and, in some cases, extra personnel might be needed to control the access and keep only a certain number of users inside the place. However, a positive point is that schemes as the limited movement policy can be implemented at any point in the pandemic and can be a key part of the measures relaxation. For instance, reducing the number of users to the minimum during the critic period of the pandemic and gradually increase this number as conditions allow. These policies may also add to avoid premature relaxation of interventions, which may produce a second wave of infections^18^, a phenomenon that has been seen in influenza pandemics^19^. Finally, these schemes can be followed when still not a vaccine is available and during the first-in-human trials^20,21^, but also be combined with vaccination strategies, once feasible.

The approaches herein developed, results and interpretations look to guide policy-makers, merchant authorities, public commercial establishments chains, and the general society to be aware of potential measures that can assist in the mitigation of the current SARS-CoV-2 pandemic, learn from them and, after implementation feedback, improve the schemes for the current and future pandemic events.

## Data Availability

No data

## Acknowledgements

We thank the support of the Alfons und Gertrud Kassel-Stiftung and the Deutsche Forschungsgemeinschaft through the project HE7707/5-1.

## Author contributions statement

G.H-M. and E.A.H-V. conceived the experiments, G.H-M. conducted the experiments, G.H-M. and E.A.H-V. analysed the results. All authors reviewed the manuscript.

